# Optimal shutdown strategies for COVID-19 with economic and mortality costs: BC as a case study

**DOI:** 10.1101/2020.11.25.20239004

**Authors:** M.T. Barlow, N.D. Marshall, R.C. Tyson

## Abstract

Decision makers with the responsibility of managing policy for the COVID-19 epidemic have faced difficult choices in balancing the competing claims of saving lives and the high economic cost of shutdowns. In this paper we formulate a model with both epidemiological and economic content to assist this decision making process. We consider two ways to handle the balance between economic costs and deaths. First, we use the statistical value of life, which in Canada is about C$7 million, to optimise over a single variable, which is the sum of the economic cost and the value of lives lost. Our second method is to calculate the Pareto optimal front when we look at the two variables – deaths and economic costs.

In both cases we find that, for most parameter values, the optimal policy is to adopt an initial shutdown level which reduces the reproduction number of the epidemic to close to 1. This level is then reduced once a vaccination program is underway. Our model also indicates that an oscillating policy of strict and mild shutdowns is less effective than a policy which maintains a moderate shutdown level.

## 1 Introduction

The COVID-19 pandemic led many countries to introduce an extensive economic and social lockdown to limit the spread of the disease (Wikipedia, 2020). While these measures are very expensive (Mandel and Veetil, 2020), in most regions where the measures were applied relatively early and with sufficient stringency, they were successful in stopping the growth of the epidemic (Moosa, 2020). After an initial period of shutdown, many jurisdictions have partially reopened their economies, and in some cases this has led to a second surge of infections (World Health Organisation, 2018) including in Canada (Grasley et al., 2020), reopening the question of whether or not to impose another lockdown.

Some kind of balance has to be struck between saving lives and the economic (and social) cost of the lockdown (Lee et al., n.d.; Miles, Stedman, and Heald, 2020). This paper presents a simple model, with both economic and epidemiological content, to help assess the options. In particular, we aim to determine what type of shutdown strategy (*X*_*t*_) minimizes costs over the period of the epidemic. The data and numbers are for the Canadian province of British Columbia. We remark that any decision on the extent of the lockdown has to take many factors into account. We have chosen to express those factors which we do consider in monetary terms; inevitably this means that some important aspects will have been left out. The optimization we perform should be viewed as information that can assist in making a complex decision, rather than as a simple prescription.

We start our model at an early stage of the epidemic, corresponding roughly to the situation in B.C. in March 2020; thus we consider ‘what should have been done’ as well as ‘what should now be done’. Moving forward, we find that the minimum cost scenario does include a significant level of economic shutdown in order to ensure reduced transmission. In particular, we obtain a high level of shutdown at the beginning of the epidemic, which is monotonically decreased until the population has been vaccinated.

## 2 Model

Our model contains two parts: One part describing the disease dynamics, the second describing the economic costs and optimal control. We separately present each part of the model below.

### 2.1 Disease Dynamics

Our model for the epidemic is of standard compartmental type. For clarity, we split it into two parts. The first part is a standard SEIQ model, where *S* is Susceptible, *E* is Exposed, *I* is infectious and *Q* is Quarantined (i.e. no longer mixing with the susceptible population while infectious):

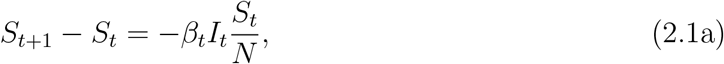

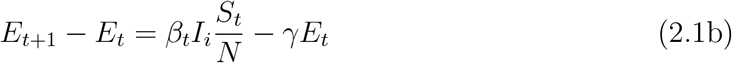

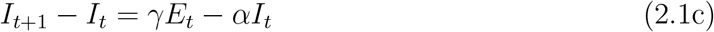

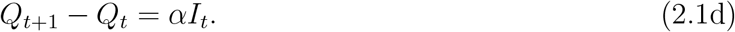

The time-dependent infectivity parameter *β*_*t*_ depends on the amount of economic lockdown, as explained below. We initialise the model by taking

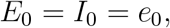

that is, the initial number of COVID-19 carriers is split evenly between the exposed and infectious compartments.

To model the progress of patients through the medical system we separate the *Q* bin into smaller bins depending on quarantine state. These bins are named *M, H, W, U, R, D*. Here *M*_*t*_ is the number of patients who are mildly ill (or asymptomatic) and therefore quarantining at home, *H*_*t*_ and *U*_*t*_ are the numbers of patients in hospital (but not ICU) and ICU beds respectively, *R*_*t*_ is the number of recovered patients, and *D*_*t*_ is the number of deceased patients. These population compartments, and the connections between them, are illustrated in Figure 1.

**Figure 1:**
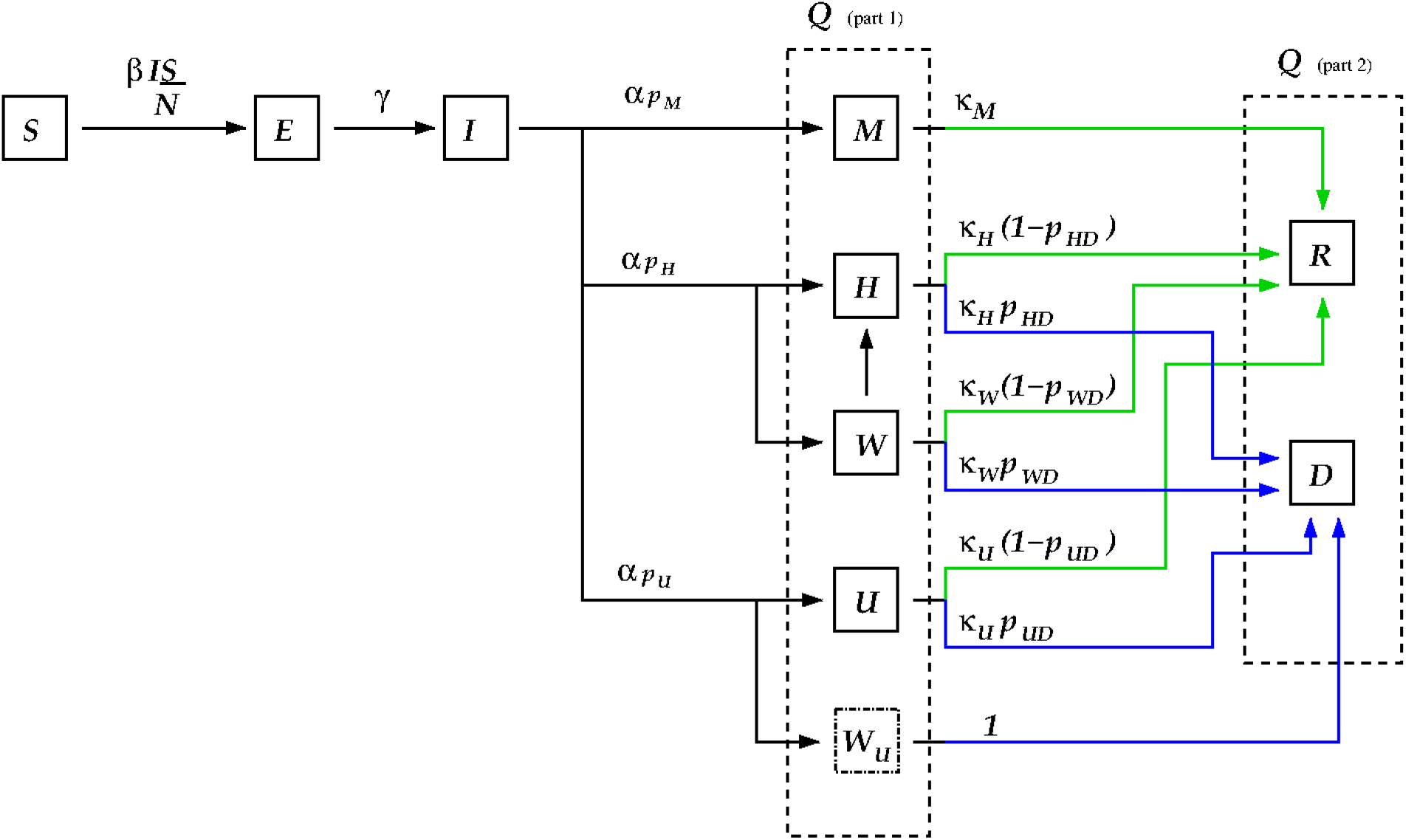
The compartments of the disease model. The two dashed line boxes surround all of those compartments that are still infectious, but are no longer transmitting disease as they are quarantined at home or in hospital (the *Q* compartment in the model). The green arrows indicate transitions from sick states to the recovered state, while blue arrows indicate transitions from sick states to the deceased state. The variables are fractions of the total population: *S* = susceptible, *E* = exposed, *I* = infectious and not quarantined, *M* = infected but only mildly ill, *H* = infected and in hospital, *W* = infected and waiting for space to open up in hospital, *U* = infected and in the ICU, *W*_*U*_ = infected and waiting for ICU space to open up, *R* = recovered, and *D* = deceased. The *W* individuals are assumed to be quarantined at home, while the *W*_*U*_ individuals transition directly into the *D* compartment.

We wish to handle possible overload of the medical system, so we set the maximum capacity of hospital and ICU beds to be *H*_*max*_ and *U*_*max*_ respectively. For patients needing a hospital (non-ICU) bed, we introduce the *W*_*t*_ compartment: The number of patients waiting for hospital beds. At time *t* + 1 the demand for hospital beds is

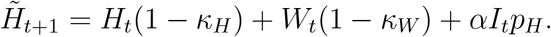

If demand is greater than supply, we move as many patients as possible into hospital, and the remainder go into the *W* container. Thus we set

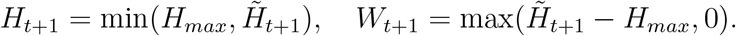

We follow a similar procedure for the ICU patients, except that (as these patients are presumably very ill) any excess is immediately moved into the *D* container; we write

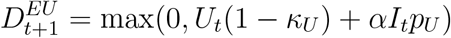

for the number of these deaths. Thus, the model for the quarantined group is written

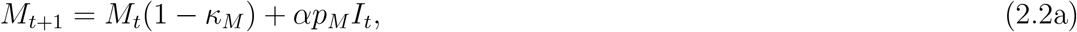

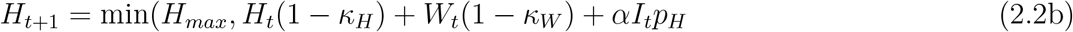

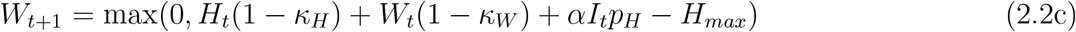

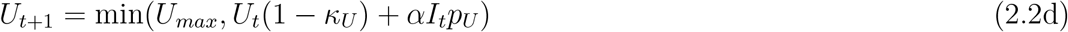

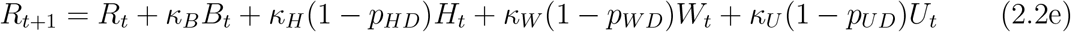

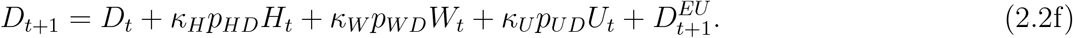

We count as *excess deaths* those deaths arising from overload of the health system. This consists of two terms: the number of patients who move into the D container due to ICU overload, together with the patients who move from *W* to *D*. We write *D*^*Excess*^ for the total number of these deaths.

Note that we have assumed that everyone who becomes infectious is eventually quarantined. While this assumption is likely unrealistically optimistic, it nevertheless means that our optimal shutdown strategy will provide a lower bound. We make similarly conservative choices with regard to the parameters contributing to the infection fatality ratio.

The disease part of the model involves a substantial number of parameters. Some of these are fairly well established but many remain quite uncertain. Table 1 summarizes the values and sources.

**Table 1:** Parameter values and sources for the disease dynamics portion of the model, equations (2.1) and (2.2). † These estimated values are in line with the rates reported in Salje et al. (2020). ‡ We were unable to find data for *p*_*W D*_, and so we assumed that survival is very low for individuals needing the ICU but unable to access those resources.

| Parameter | Value | Remarks |
| --- | --- | --- |
| $\alpha$ | .17 | Anderson et al. (2020) |
| $\gamma$ | 0.2 | Anderson et al. (2020) |
| $\mathcal{R}_0$ | 3.3 | Salje et al. (2020) |
| $\beta$ | $\alpha\mathcal{R}_0$ | Derived |
| $\kappa_M^{-1}$ | 14 | estimated† |
| $\kappa_H^{-1}$ | 10 | Salje et al. (2020) |
| $\kappa_U^{-1}$ | 15 | Salje et al. (2020) |
| $\kappa_W^{-1}$ | 2 | estimated† |
| $p_H$ | 0.021 | Salje et al. (2020) |
| $p_U$ | 0.0047 | Salje et al. (2020) |
| $p_M$ | $1 - p_H - p_U$ | Derived |
| $p_{HD}$ | 0.17 | PHAC Emerging Sciences (2020) |
| $p_{UD}$ | 0.27 | PHAC Emerging Sciences (2020) |
| $p_{WD}$ | 0.8 | estimated‡ |
| $e_0$ | 200 | Initial values of $I_0$ and $E_0$ |

With these parameter values the total infection fatality rate (IFR) without overload is

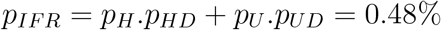

which is at the low end of the range of values reported in the literature (PHAC Emerging Sciences, 2020).

### 2.2 Economic Costs

We now describe the economic/control part of the model. The proportion of the economy which is shut down (SD) on day *t* is given by *X*_*t*_*∈*[0, *x*_*m*_]. Here *x*_*m*_ < 1, as parts of the economy, e.g. food distribution, cannot be shut down. We assume that the shutdown is arranged so that the parts of the economy with the biggest effect on *β* are chosen first; thus we obtain a concave effectiveness from the shutdown.

Let *f* (*x*) be the proportional reduction in social contacts due to the shutdown; we call *f* the *shutdown effectiveness* function. A shutdown of *X*_*t*_ will lead to an infection rate on day *t* of

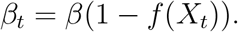

We do not have the data to calculate *f* in detail. A simple model is to take

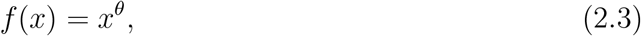

(see Figure 2) where *θ* < 1. The reproduction number due to a SD of *x* is then given by

**Figure 2:**
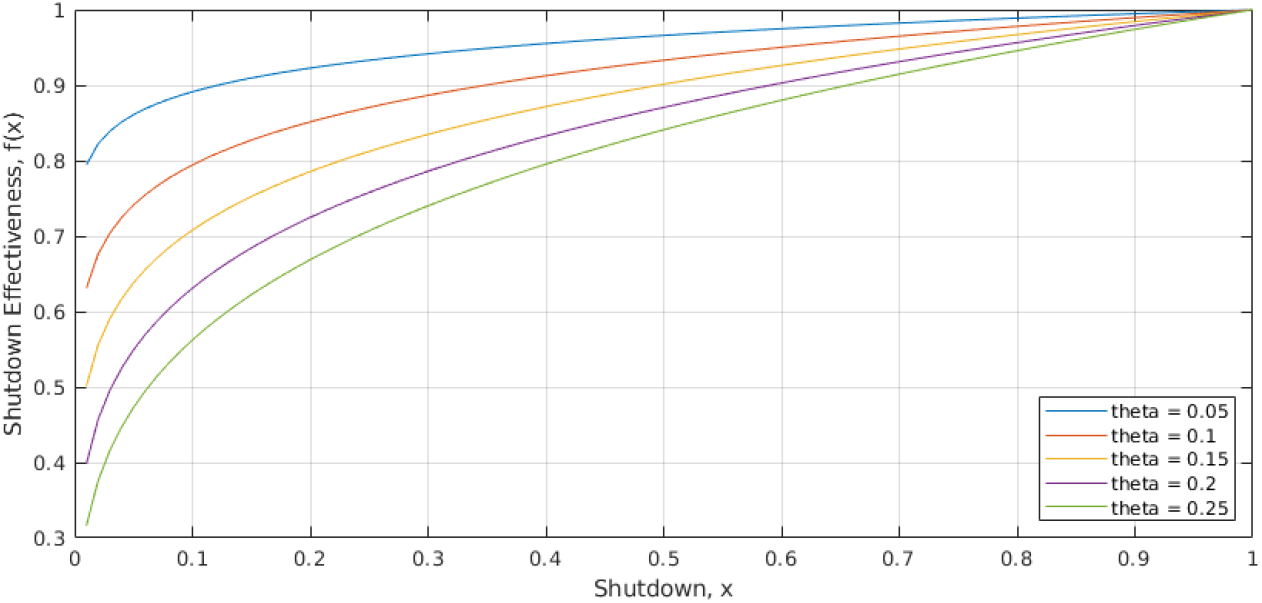
The shutdown effectiveness function, *f* (*x*) = *x*^*θ*^. The higher the value of *θ*, the greater is the reduction in effective reproduction number for a given level *x* of economic shutdown.

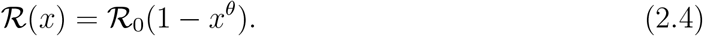

We note that *d*ℛ_0_(0+)*/dx* = −∞, which implies that some level of shutdown is always advantageous – see Appendix B.

With an effectiveness function of this form we need to estimate *θ*. A first guess would be to use the ‘Pareto principle’, which states roughly that “80% of the output is due to 20% of the input”. We thus write *f* (1*/*5) = 4*/*5 which gives *θ*≈0.14. This guess has some support from data: The fall in GDP in Canada in the first quarter of 2020 is estimated to lie between 7% and 14% (Infoline, 2020a,b; Statistics Canada, n.d.). Anderson et al. (2020, page10) estimates that the March-April 2020 shutdown in B.C. reduced contacts by about 78%, with a 90% credible interval of 66% to 89%, which reduced ℛ (*x*) to below 1. Using these values for GDP and contact reduction in (2.3), we obtain *θ∈*[0.04, 0.21]. The highest values of *θ* are obtained for the highest fall in GDP, and the lowest reduction in contacts. We note that the GDP estimates do not include capital destruction costs, which tend to take longer to become evident (Erken et al., 2020), and so we assume that the ultimate cost is actually higher than estimated. It therefore seems prudent to set *θ* at a value closer to the upper end of the plausible range. We thus select *θ* = 0.2 as the default value.

We write *x*_*c*_ for the amount of SD needed to make ℛ equal to 1; thus

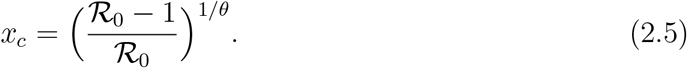

If ℛ_0_ = 3.3 and *θ* = 0.2, then *x*_*c*_ = 0.164.

The equations above describe the evolution of the epidemic for the period *t* = 0, 1, …, *T*_*V*_ once we have fixed the initial conditions and the control *X*_*t*_. We take *T*_*V*_ to be 360 days; this is the approximate time from the start of the epidemic (March 2020) until we might hope that a vaccine is available. Once a vaccine is available, supply constraints mean that it will still not be possible to vaccinate the whole population of B.C. all at once, and so some level of shutdown may still be necessary for *t* > *T*_*V*_. We assume that the vaccine is 100% effective and that *N*_*V*_ people are vaccinated daily for each day *t* ≥ *T*_*V*_. For each day *t* ≥ *T*_*V*_ we move *N*_*V*_ people from *S* to *R*, and otherwise continue to run the model as above. We take a terminal time *T* = *T*_*V*_ + *N/N*_*V*_ for the model; at this point the whole population is vaccinated and the epidemic is at an end. In fact, herd immunity will be reached before the terminal time *T* (Garibaldi, E.R.Moen, and Pissarides, 2020), but some costs are likely to extend past this point, and so we take the larger terminal time as our end time. With our base parameter values we have *T*_*V*_ = 360 and *T* = 610.

We now introduce costs; throughout the paper these are in Canadian dollars. The costs we consider are the economic costs of the shutdown *C*^(*SD*)^, costs arising from the medical care of infected individuals *C*^(^*M*), and costs due to lives lost *C*^(^*D*).

The economic cost of shutdown is taken to be the daily shutdown costs summed over the time period of the the epidemic, so that writing *G*_*D*_ to for the daily GDP of B.C. we have

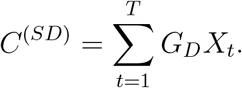

Medical costs can be inferred from Jones (2020); we take the cost of a day in a regular hospital bed as $1,000, and ICU costs as $10,000 on the first day and $2,000 per day thereafter. The medical cost during the period *t* = 0, …, *T* is therefore

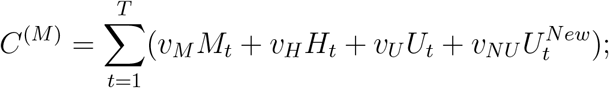

here *v*_*M*_, *v*_*H*_, *v*_*U*_ are the daily costs for patients in states *M, H* or *U, v*_*NU*_ is the excess cost for the first day in ICU, and 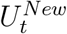 is the number of new patients in ICU on day *t*.

To determine costs due to the lives lost, we need to determine the ‘statistical (dollar) value of a life’ (SVL) or *V*_*L*_. There is a substantial literature on this topic – see Dionne and Lanoie (2004), Greenstone and Nigam (2020), and Thunström et al. (2020); authors use a variety of methods to get some handle on this number. As one would expect, estimates vary widely – the review in Dionne and Lanoie (2004) gives a range for Canada of C$2.0m – C$11.1m, with a median of C$5.5m. In 2020 dollars that is *V*_*L*_ = *C*$7.0m, which we take as our base figure. We then set

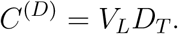

The total cost of the epidemic is the sum of the shutdown, medical, and death costs:

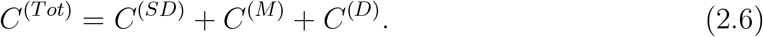

Note that the first two of the costs in (2.6) are pure dollar costs, while the third is based on a more subjective evaluation of the dollar cost of a lost life. We use the SVL in order to estimate the total cost of the epidemic, but since the quantitative assessment of the pure dollar and SVL costs are so different, in our analysis below we also treat these two types of costs separately. We write

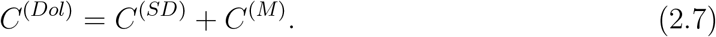

We list the economic parameters in Table 2.

**Table 2:** Economic parameters. Dollar values are in Canadian dollars. For the lost wages per day of illness, *v*_*M*_, we take the drop in GDP and divided by the number of days over which the drop occurred. For the number of daily vaccinations after *T*_*V*_, we take the population of Canada (Statistics Canada, 2020) divided by the estimated length of time it will take to vaccinate everyone (Miller and Zafar, 2020).

| Parameter | Value | Remarks |
| --- | --- | --- |
| $N$ | 5,100,000 | Population of BC (Statistics Canada, 2020) |
| $G_D$ | \$800,000,000 | daily GDP in B.C. (Statistics Canada, n.d.) |
| $v_M$ | \$100 | Lost wages per day of illness |
| $v_H$ | \$1000 | Day in hospital (Jones, 2020) |
| $v_U$ | \$2000 | Day in ICU (Jones, 2020) |
| $v_{NU}$ | \$8000 | ICU setup (Jones, 2020) |
| $V_L$ | \$7,000,000 | Value of life (Dionne and Lanoie, 2002) |
| $T_V$ | 360 | Number of days until vaccination starts |
| $N_V$ | 20400 | Number of daily vaccinations after $T_V$ |
| $T$ | $T_V + N/N_V$ | Terminal time for model |
| $\theta$ | 0.2 | Effectiveness of shutdown: range 0.1 – 0.2. |

### 2.3 Optimization procedure

We considered two types of optimization. In section 3.1 we minimized the total cost of the epidemic using value of life *V*_*L*_ to translate lost lives into costs. Since it does not always make sense to translate a lost life into a dollar value, we take a different approach in Section 3.2. There we looked at Pareto optimization (Mock, 2011) of pure dollar costs and deaths, i.e. for the two numbers (*C*^(*Dol*)^, *D*_*T*_). We studied the structure of the epidemic associated with some sample points on the Pareto curve.

In both cases we only considered strategies in which *X*_*t*_ was held constant for a significant time period. For the total cost optimization we divided the time period into 10 equal segments of 61 days (about 2 months), and held *X*_*t*_ constant in each of these periods. Writing *x*_*i*_ for the value of *X*_*t*_ in period *i*, the strategy (*X*_*t*_) is described by the vector *x* = (*x*_1_, …, *x*_10_). We used Matlabs’s multivariate optimization routine “fmincon” to optimize over vectors *x*. For the Pareto optimization we used 5 equal time periods, each of 122 days (the Pareto optimisation performed poorly with 10 periods).

We had several reasons for this restriction on shutdown strategies. First, from a computational point of view, it is not feasible to optimize over several hundred control values *X*_*i*_. Next, since the effectiveness function *f* is concave, rapidly varying control strategies will perform less well than more slowly varying control strategies (see Appendix A). Finally, as it is not realistic to consider that a government could implement a shutdown function *X*_*t*_ that changes too frequently.

We remark that it would be as computationally feasible to consider piecewise linear strategies as piecewise constant ones – in both cases the optimization reduces to optimizing over a 10-dimensional space (if we use our baseline setup). However, we did not consider such strategies because we did not consider them to be applicable in practice.

The two types of optimization are connected, since the total cost optimization corresponds to tangents on the Pareto curve. More precisely, write (*C, D*) for the two axes (dollar costs and deaths) for the Pareto curve. Let *vD* + *C* = *b* be a tangent to the Pareto curve at the point (*C*_1_, *D*_1_). Then *b* = *C*_1_ + *vD*_1_ is also the minimum total cost when we take *V*_*L*_ = *v*.

## 3 Results

Before we consider the optimization problem in detail, it is helpful to look at the basic structure of the model described above. If the epidemic is such that there is no overload of hospital capacity, and so no excess deaths, then the total number of infected individuals is *R*_*T*_ + *D*_*T*_, and the total number of deaths is

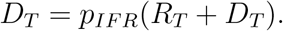

We can compute the expected ‘value of life’ costs as the probability of dying times the value of a lost life, i.e. *V*_*L*_ *p*_*IFR*_ = $33, 600, for our baseline parameter values. On the other hand, the expected medical costs are

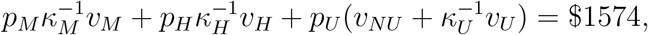

for the baseline parameter values. So, on average, the medical costs of the epidemic are small in comparison with the value of life costs.

### 3.1 Optimization of total costs

#### 3.1.1 Baseline (constant shut down) scenarios

The simplest strategy is to take *X*_*t*_ = *x* constant for the whole period. Figure 3 shows the total cost as a function of *x* for various values of the SVL *V*_*L*_. This graph has several informative features. We consider first the curves with *V*_*L*_ ≥ $3.0m. In these cases, the minimum value *x*_*min*_ is a little smaller than *x*_*c*_ = 0.164 (if *V*_*L*_ = $7.0m then *x*_*min*_ = 0.13). These values of *x* correspond to allowing the epidemic to grow slowly over the pre-vaccination period; it then declines once vaccination reduces the effective reproduction rate. One sees further that the minimizing value *x*_*min*_ is not very sensitive to *V*_*L*_. These values of *x*_*min*_ give rise to a relatively small epidemic, so any further increase in *x* would save relatively few lives. For *x* > *x*_*c*_ the total cost curves for differing values of *V*_*L*_ are very close together; this is because in this regime the total number of deaths is small (a few hundred), so the term *V*_*L*_*D*_*T*_ is much smaller than the pure dollar cost *C*^*Dol*^.

**Figure 3:**
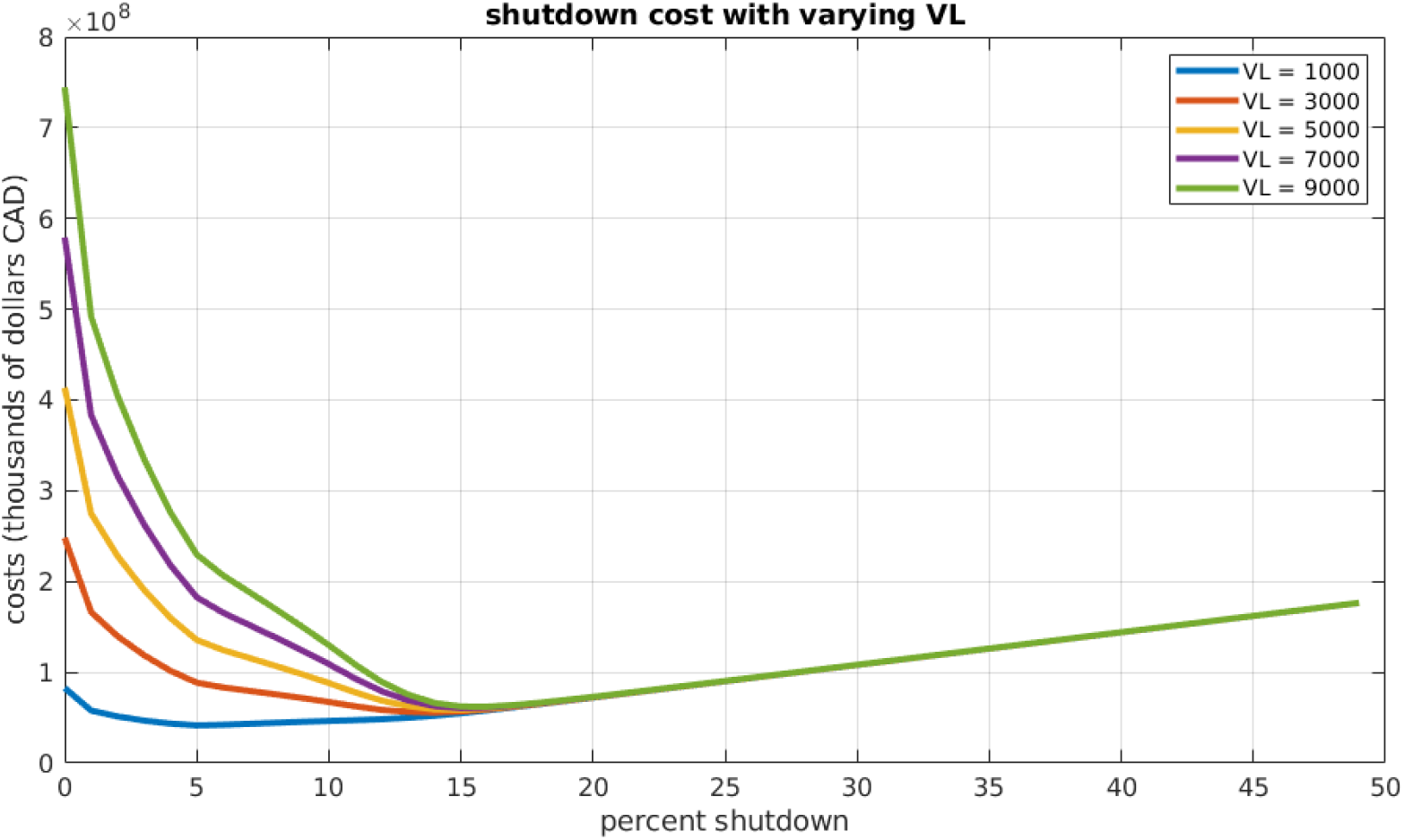
Total costs as a function of shutdown level for a constant shutdown strategy. Several values of *V*_*L*_ (in thousands of dollars CAD) are shown.

For *V*_*L*_ = 1.0m one sees a rather different pattern. The minimum cost is obtained at a much lower shutdown level, around 5%. In this case, deaths are counted as relatively unimportant, so the optimal strategy is to adopt a relatively mild shutdown.

#### 3.1.2 Variable shutdown scenarios

We next consider strategies that are piecewise constant, being allowed to vary only every 61 days. We see the same general pattern as with a constant shutdown. For the baseline parameter values we find that the optimum policy (Figure 4, left plots) starts with a fairly strict shutdown, with *X*_*t*_ ≈ *x*_*c*_ = 0.164, for the first six time periods. Subsequently, *X*_*t*_ is gradually relaxed, and is zero in the final period. Taking *X*_*t*_ < *x*_*c*_ means that the epidemic grows over the first year, but at a slow and controlled rate. As vaccinations proceed, the optimum policy is to relax the shutdown enough to allow a late peak in the epidemic. This peak occurs during the period when the vaccine has become available. If the initial infection level is ten times higher than the baseline (Figure 4, right plots), we see a similar pattern, but with higher shutdown levels at the beginning, and shutdown levels dropping more in the first third of the year. Notice that in this second scenario, the optimal strategy designates *X*_*t*_ > *x*_*c*_ initially, so that the epidemic decreases, bringing the case load under control before resuming the late wave pattern of the baseline case.

**Figure 4:**
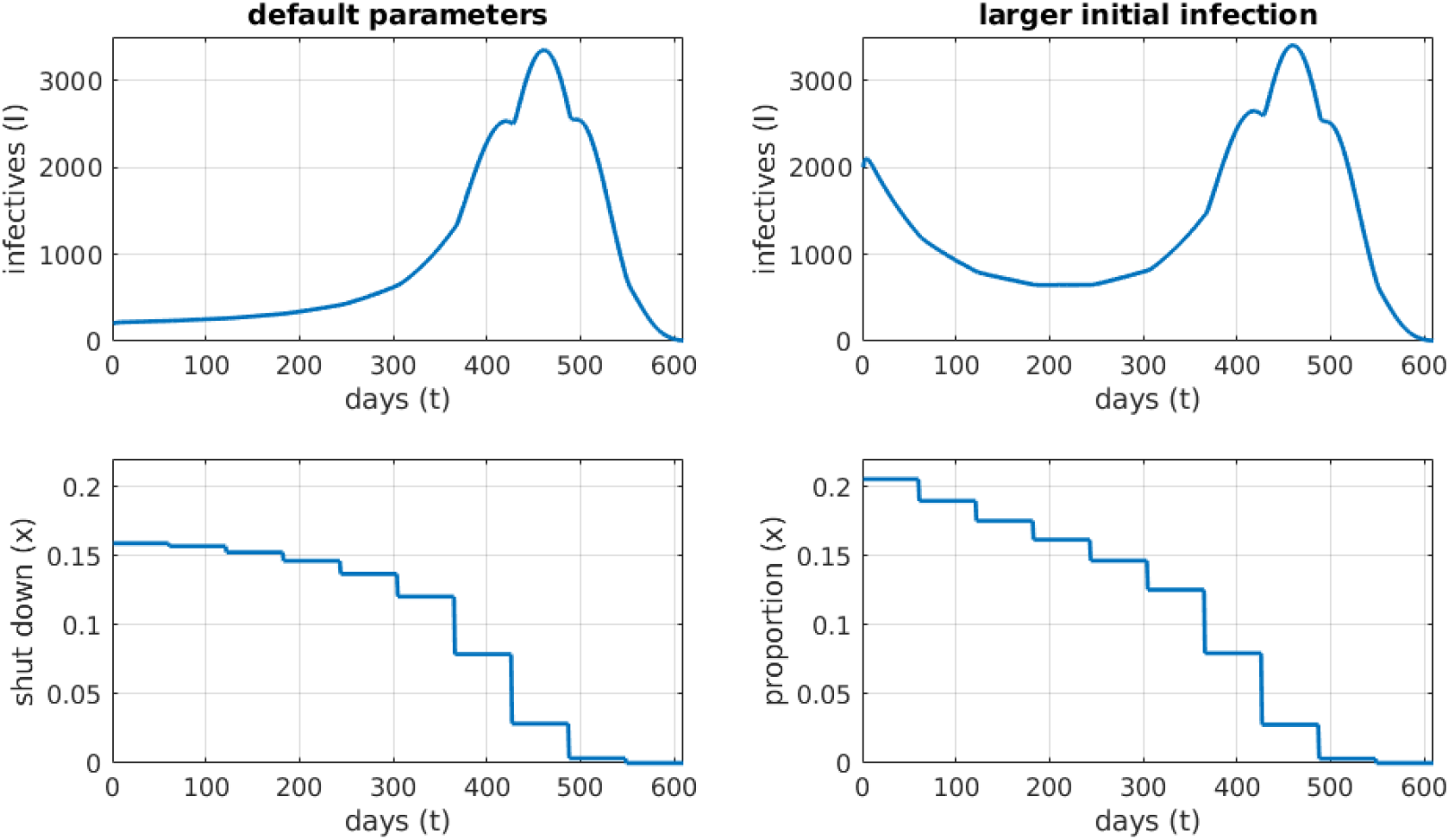
Number of infections and shutdown for optimal policy for base parameters (left) and a larger initial epidemic with *I*_0_ = *E*_0_ = 2000 (right) Need bigger axis labels!, minimizing total costs, *C*^(^*Tot*) (see (2.6)). For these parameter values, the critical shutdown level is *X*_*c*_ = 0.164. Vaccination begins at *T*_*V*_ = 360.

The overall pattern can be characterised as a high level of shutdown at the beginning of the epidemic, with *X*_*t*_ decreasing in small steps at first, and then in larger steps as a vaccine becomes available. The change in *X*_*t*_ is monotonic: Under optimal conditions, there is no second wave. This pattern held for most of the runs we performed with modified parameters: see Table 3 for a summary of the outcomes. Increasing the number of time periods to 20 makes little difference to the total cost or number of deaths.

**Table 3:** Economic and epidemiological outcomes corresponding to the optimal solution of the model under variation of a few key parameters (left column). The Total Cost outcome includes the value of lives lost (all dollar values are in CAD). For each row in the table, all parameters except the one being varied are held at the default values listed in Tables 1 and 2. For comparison, the outcomes under default values for all of the parameters are listed in the first row.

| Parameter varied | Total Cost (\$billion) | Deaths | Max infected | Excess deaths |
| --- | --- | --- | --- | --- |
| None | 51.4 | 494 | 3,345 | 0 |
| $V_L = \$5.0\text{m}$ | 50.3 | 697 | 4,783 | 0 |
| $V_L = \$3.0\text{m}$ | 48.4 | 1,228 | 8,435 | 0 |
| $V_L = \$2.0\text{m}$ | 46.9 | 2,214 | 13,835 | 0 |
| $V_L = \$1.0\text{m}$ | 34.1 | 20,477 | 161,167 | 8,751 |
| $V_L = \$0.0\text{m}$ | 6.8 | 64,339 | 584,296 | 55,932 |
| $\theta = 0.15$ | 27.8 | 347 | 2,124 | 0 |
| $\mathcal{R}_0 = 3.6$ | 62.5 | 532 | 3,657 | 0 |
| $\mathcal{R}_0 = 3.0$ | 40.5 | 445 | 2,936 | 0 |
| $e_0 = 2,000$ | 59.0 | 665 | 3,408 | 0 |
| $e_0 = 20,000$ | 68.6 | 1,250 | 20,782 | 0 |
| 20 time periods | 51.3 | 494 | 3,105 | 0 |
| 1 time period | 69.6 | 811 | 5341 | 0 |

As in the constant policy case, for *V*_*L*_ ≤ $1.0m the optimal strategy changes to one which permits a large epidemic, with many deaths. The shutdown values for the first three periods are 0.0001, 0.0684, 0.0718. One might expect that a strategy which is the same for the final 7 periods, but is the average of the first three at the beginning (i.e. 0.0467), would perform better, but this is not the case. This modification to the optimum strategy delays the epidemic peak, but gives less social distancing when it is needed most, and allows a bigger epidemic, with 31,010 deaths as opposed to 20,477. See Figure 5.

**Figure 5:**
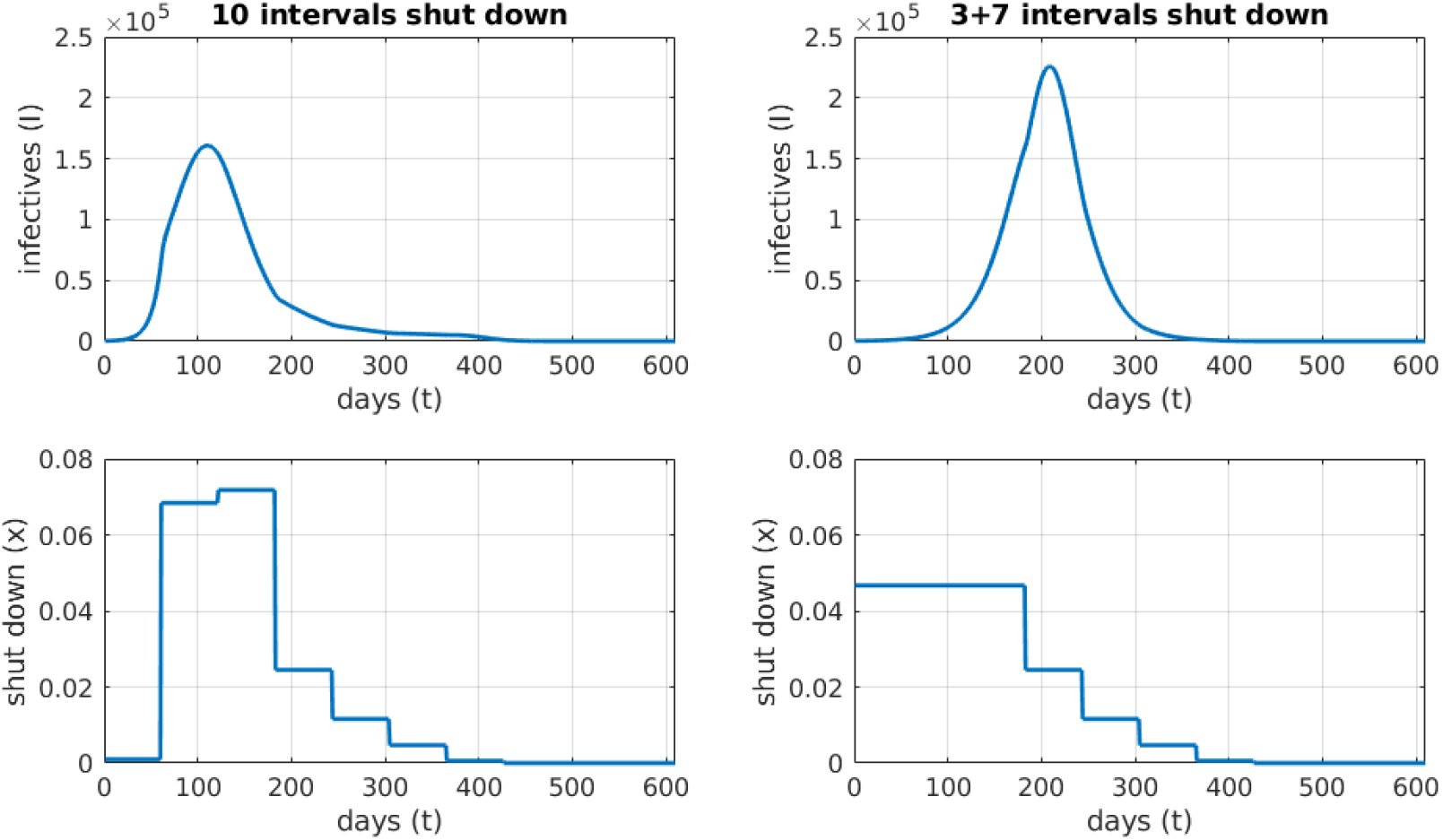
Effect of a lower *V*_*L*_ = $1.0m. Number of infections and shutdown for the optimal policy (left), and a modified control held constant at the average value of the optimal control for the first three time periods (right). The number of deaths for the optimal control is 20,477, and for the modified control is 31,010.

**Figure 6:**
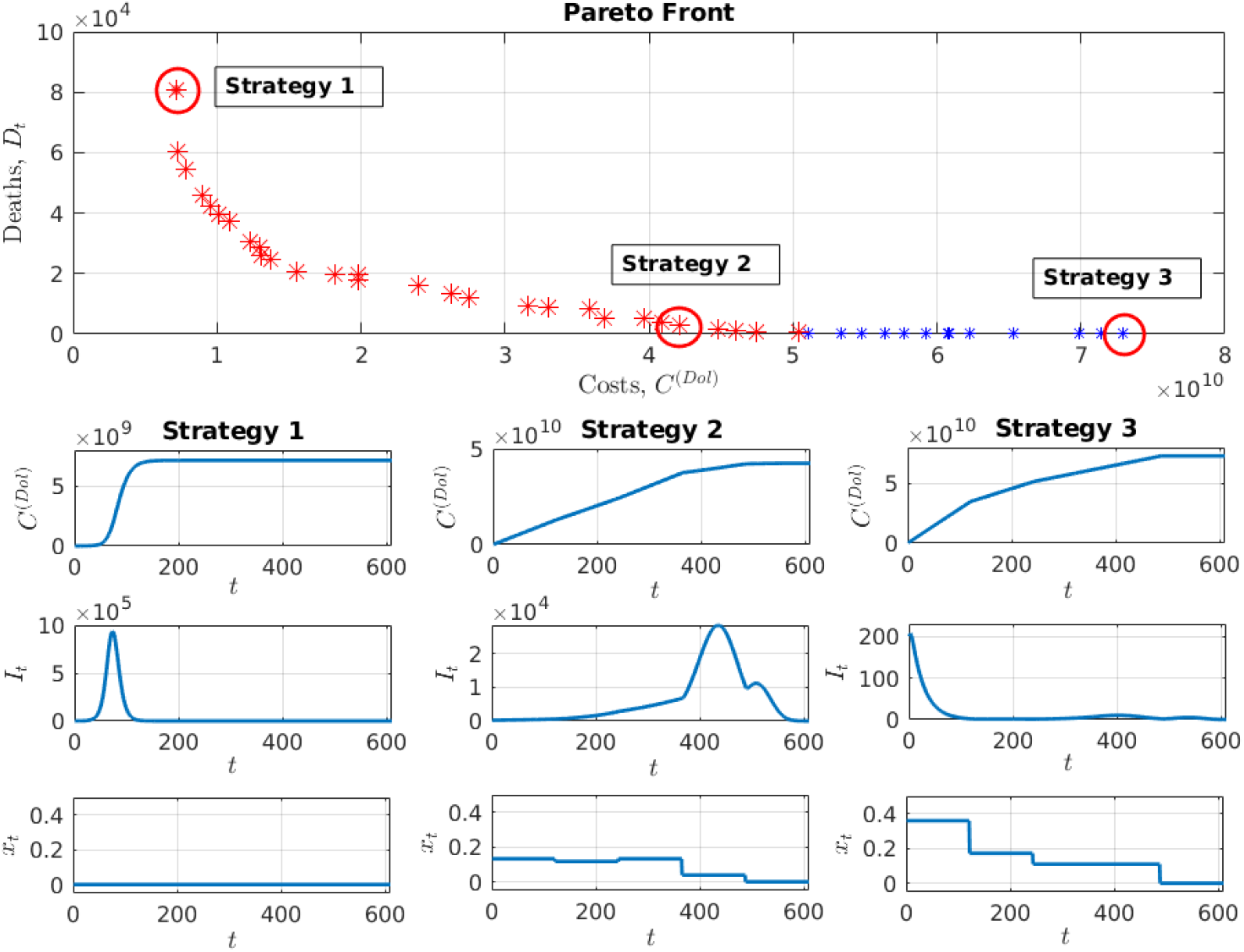
Top: Pareto plot. Each asterisk corresponds to the shutdown strategy which gives the minimum possible number of deaths *D*_*t*_ at dollar cost *C*^(*Dol*)^ (recall that dollar cost does not include the value of a lost life). The Pareto front assumes default parameter values and five shutdown intervals with possibly varying levels of social distancing. Points on the Pareto front with excess deaths are red and those without are blue. The three highlighted strategies are: Do nothing (minimize costs, excluding value of life), do a some of both (try to minimize deaths and costs simultaneously), and implement severe distancing (minimize deaths). Bottom grid of plots: The cumulative cost (top row), daily infections (middle row), and daily shutdown strategy (bottom row), are shown for the epidemics associated with the three points circled in the Pareto plot (Strategies 1-3). Note that the shutdown strategy for Strategy 1 is zero (the vertical axis includes negative values - for this subplot only - so that the zero strategy line is visible.

Thus, apart from the need to control a large initial epidemic, one finds one wants *X*_*t*_ close to *x*_*c*_ in the pre-vaccination phase. Taking *X*_*t*_ significantly smaller than *x*_*c*_ causes a large epidemic, with many deaths, while taking *X*_*t*_ significantly larger than *x*_*c*_ is economically costly, and saves few lives.

### 3.2 Pareto optimization: Costs and Deaths

In the results presented above, we find the optimum shutdown pattern by expressing all of the constraints, including lives lost, in terms of their monetary value, *C*^(*Tot*)^. Here, we separate out pure dollar costs *C*^(*Dol*)^ from deaths, applying the two as separate constraints, and present the Pareto optimum (Mock, 2011). The results are shown in Figure 3.2. The curve shows the competing effects of minimizing costs versus minimizing deaths: Minimizing the pure dollar costs of shutdown leads to higher mortality, and minimizing mortality leads to higher pure dollar costs.

Each point (*x, y*) on the Pareto curve corresponds to the shutdown strategy which gives the minimum possible number of deaths *y* at pure dollar cost *x*. Three of these Pareto-optimum shutdown strategies are shown in the lower half of Figure 3.2, and the Pareto points they correspond to are circled on the Pareto curve. The extreme strategies are: (Strategy 1) prioritise minimizing pure dollar costs over deaths, and (Strategy 3) prioritize minimizing deaths over costs. We also show an intermediate strategy (Strategy 2), where costs are half that of Strategy 3, but mortality is still close to the minimum.

If minimizing pure dollar costs is the highest priority (Strategy 1), then the optimal strategy is to have no economic shutdown at all, and we obtain a death toll of over 80,000 individuals. The total cost is less than $10 billion. If minimizing deaths is the highest priority (Strategy 3), we obtain a shutdown strategy with an initially high shutdown level, which then decreases at each time period until finally becoming zero at the last interval. The cost skyrockets to over $70 billion, and the number of deaths is less than 100. If, instead, we choose the intermediate strategy (Strategy 2), pure dollar costs can be reduced to near half that of the most expensive strategy, and the total deaths are a few thousand. Table 3 lists economic and epidemiology outcomes for a few additional scenarios. These results provide context to the Pareto curve in Figure 3.2. We do not see excess deaths (and a large decrease in cost) until *V*_*L*_ drops to $1.0m or less. Increasing the frequency with which the control strategy is adjusted (i.e., increasing the number of time periods) has very little effect on any of the outcomes. In contrast, decreasing the adjustment frequency down to the point where only one constant shutdown level is used throughout the epidemic results in almost twice as many deaths and actually increases the total cost. Decreasing the shutdown efficiency *θ* and basic reproduction number ℛ_0_ both lead to reduced cost and deaths, while increasing ℛ_0_ and the initial disease prevalence *e*_0_ both lead to increased costs and deaths.

### 3.3 Sensitivity Analysis

Since some of the parameters are not well known, we test our results using a Partial Rank Correlation Coefficient (PRCC) sensitivity analysis as described by Marino et al. (2008). PRCC is a reliable measure of the contribution that parameters have to the model when the relationship between the parameter and the output is monotonic. Each parameter is varied over a range from half to twice the baseline value given in Tables 1 and 2. The PRCC values can be interpreted as the correlation between parameter and model output, linearly discounting the effects of the other parameters.

We ran the sensitivity analysis on the model with the optimal strategy given for the baseline parameters, and considered the sensitivity (a) with respect to both total costs (i.e. the sum of pure dollar and value of life costs), and (b) with respect to deaths. Both sensitivity analyses show high (greater than 0.6) sensitivity values for the parameters *R*_0_ and *θ*. In addition, for costs *T*_*V*_ and *N*_*V*_ had moderate sensitivity, in the range (0.2 to 0.6). For deaths, the parameters *pH, pHD, pUD* and *e*_0_ had moderate sensitivity. See the supplementary material for full details. In both cases the parameters 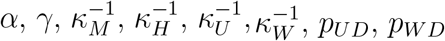 had low sensitivity – less than 0.11.

It may seem somewhat surprising that *V*_*L*_ has a relatively low sensitivity of 0.19. One reason for this is that our sensitivity analysis only studies sensitivity of a parameter *x* over the range [*x*_0_*/*2, 2*x*_0_], where *x*_0_ is our base value. Thus for *V*_*L*_, the sensitivity analysis only considered the range between $3.5m and $14m, and, as we already saw when we looked at constant shutdowns, in this range the value of life makes little difference to the strategy or to the total cost.

## 4 Discussion

Our study is motivated by the hesitation shown by some leaders to implementing strict control measures to slow the spread of COVID-19 (Haberman and Senger, 2020; Mandl and Benassatto, 2020; Maynes, 2020), in part due to the huge toll to the economy. For informed decision making, it is clear that we need some objective quantification of the total cost of both the health crisis and the economic shutdown measures. In this work, we present a coupled disease and economic cost model which is a useful tool for evaluating shutdown options. While our model is in no way a comprehensive representation of all of the costs and benefits of shutdown measures, we submit that it contains the salient features of the system, and so the patterns in our results should reflect real dynamics.

For a simple minimization it is necessary to combine our two key variables, that is the dollar cost (medical costs plus economic shutdown costs), and deaths, which we did by using the ‘value of a statistical life’ *V*_*L*_. For values of *V*_*L*_ close to the consensus value of about $7.0m given in Dionne and Lanoie (2004) our results indicate that total costs (deaths, hospital costs, and deaths costs) are minimized by a significant shutdown – of about 10%-15% in the early stages of the epidemic. For these values of *V*_*L*_ the optimal shutdown strategy decreases slowly in the initial phases of the epidemic, and then falls rapidly to nearly zero once vaccination is well underway and the system is close to herd immunity. In the middle of the vaccination phase the shutdown is relaxed sufficiently to allow a moderate late peak in infections. At this point the effective reproduction number has been reduced sufficiently so that the epidemic has no possibility to explode before the completion of the vaccination program.

If *V*_*L*_ is taken to be $1m or less, a different pattern occurs. As death costs are smaller, economic costs play a relatively larger role, and the question is how best to deploy the relatively small amount of shutdown that will be introduced. It turns out that the optimal control is to wait until the infections are rising rapidly, and then use the shutdown to reduce the height of the epidemic peak.

In this paper we have studied the optimization problem for the situation when the disease and economic parameters are known. In practice of course this is not the case, and in particular neither of the two most significant parameters (ℛ_0_ and *θ*) would have been known to a government considering lockdowns. One therefore needs to consider strategies using feedback to control the epidemic. In the context of our model, a reasonable goal would be to aim at having the effective reproduction number 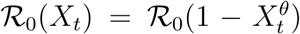 close to 1. The results of Stewart, Van Heusden, and Dumont (2020) suggest this goal is reasonably attainable.

### 4.1 Gradual versus periodic shutdowns

Our observations provide an interesting perspective on the shutdown approaches taken by governments in BC and elsewhere. In almost all cases, after a severe initial shutdown, economies were reopened and it was hoped the transmission rate could be controlled through contact tracing as well as individual prophylactic behaviours (mask-wearing, washing hands, avoiding crowds, keeping business patrons 2 m (6’) apart, etc.). Unfortunately, these methods do not seem to be as effective as was hoped, and as we write (November 2020) cases are rising dramatically in many jurisdictions, and governments are reimplementing significant shutdown measures (Wikipedia, 2020). From our work, it appears that a slower and more gradual decrease in shutdown level would have led to a smaller overall cost of the epidemic.

### 4.2 Cost of lives lost

Since the equating of a life lost to a dollar value is debatable, we found it useful to separate pure dollar costs and deaths. The Pareto curve shows us that deaths can be maintained at a very low level, even if the shutdown level falls short of “stop the epidemic”. Again, unless it is acceptable to let the number of deaths skyrocket, the optimal shutdown strategy always includes a positive and significant (i.e. over ≈15%) level of shutdown.

In this work we use a single value for the value of life *V*_*L*_: that is, we do not make any adjustments for the age of the deceased. If we neglect excess deaths, then this gives a SVL cost of *C*_0_ = *p*_*IFR*_*V*_*L*_ per infection; with our base parameters this is $36,400. It is natural to ask what happens if we look at an age stratified population. Mortality rates are substantially higher for older patients Salje et al. (2020), and as they have a smaller life expectancy it can be argued that their SVL should be smaller.

To assess this point, we can look at the following simple age stratified model. We divide the population into *n* age groups. Group *i* consists of a proportion *a*_*i*_ of the population, has death rate (IFR) *p*_*i*_ and has SVL *v*_*i*_. Then if we neglect excess deaths, the overall cost per infection is

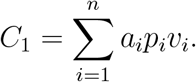

Let *v*_*i*_ for each age group be the life expectancy of the mid point of the age range times $170,000, which following we take to be the value of a year of life. Using the age-related death rates from Salje et al. (ibid.) and BC census data from 2011, we obtain *C*_1_ = $12, 377, i.e. about 38% of *C*_0_ = *p*_*IFR*_*V*_*L*_=$33,600. Thus the age stratified model outlined above corresponds to taking *V*_*L*_ to be 37% of the value listed in Table 2, i.e. about $2,600,000. (See the supplementary information for more details.) The outcomes listed in Table 3 show that for *V*_*L*_ as low as $2.0 m, the optimal shutdown strategy is sufficiently large to prevent any excess deaths, and so, broadly speaking, our results are not changed. Note that this calculation neglects excess deaths, and assumes that all the disease parameters except death rates are independent of age.

### 4.3 Other costs

Since our focus is the epidemic in BC, we make a few remarks about the specific situation there, namely, the overdose crisis. While our work strongly suggests that the adoption of economic shutdown measures in BC is, in the long term, a responsible strategy, the shutdown in BC has caused a significant increase in deaths due to drug overdoses there (BC Coroners Service, 2020); the excess (total overdose deaths over and above the usual average) for March through June 2020, exceeds the total number of COVID-19 deaths for that time period by over 40% (BC Centre for Disease Control, 2020; BC Coroners Service, 2020). A number of factors are likely involved in this mortality, including the disruption of regular drug supply chains, through border closures, leading to an increasingly toxic drug supply (Zussman, 2020), the reduction in access to harm reduction services as a result of physical distancing protocols (ibid.), and increased stress resulting from increased isolation and economic uncertainty (Ebrahimi, Hoffart, and Johnson, 2020; Hyland et al., 2020; Lechner et al., 2020) (similar patterns have been observed in the US (Katz, Goodnough, and Sanger-Katz, 2020)). The value of these lost lives could have a significant effect on our cost calculations, but assessing the relation between the level of economic shutdown and the number of overdose deaths is not straightforward. The social issues around the legality of drugs, addictions treatment, and overdose deaths is well beyond the scope of this paper, but is nevertheless an extremely important issue.

There are many additional costs to social distancing that have not been included in our model due to lack of data. We discuss some of the most important of these here. First, while our economic cost calculations take account of loss of wages and business income due to the shutdown, it does not include costs due to capital destruction. The Canadian Federation of Independent Business recently estimated that 12% of small and medium-sized BC businesses are at risk of closure (Gaudreault, 2020; McCusker and Tindale, 2020), but estimates of the effect of these possible closures on provincial GDP is not yet known. Second, with hospitals stretched to accommodate COVID-19 patients, and with fear of contracting COVID-19 limiting movement, treatment of non-COVID illnesses can be severely delayed (Dayal et al., 2020; Lazzerini et al., 2020). Thus there will be excess deaths from other causes, and costs associated with suffering due to untreated conditions (Dayal et al., 2020; Solis et al., 2020). Finally, mental health, in general, is known to suffer during community disasters (Lowe et al., 2019; North and Pfefferbaum, 2013), including pandemics. Research is emerging regarding the effects of COVID-19 non-pharmaceutical measures on mental health (Ebrahimi, Hoffart, and Johnson, 2020; Hyland et al., 2020; Lechner et al., 2020; Van Rheenen et al., 2020; Vinkers et al., 2020), but costs are difficult to determine at this stage.

### 4.4 Future considerations

Our understanding of the SARS-Cov-2 virus and COVID-19 disease are continually evolving, and so necessarily our model is relevant to the current epidemic up to a certain time point. In particular, we do not consider the possibility of a successful treatment that reduces mortality among those who become infected. There is evidence that treatment outcomes are improving for COVID-19 patients (National Institute of Health (USA), 2020), but the data are still preliminary and therefore difficult to include. We also do not include contact tracing as a measure for reducing transmission. Considerable work evaluating the effectiveness of contact tracing has been done by other researchers (Firth et al., 2020; Kucharski et al., 2020), and the general consensus is that contact tracing alone is insufficient, and can quickly become overwhelmed by a few large-scale outbreaks.

Finally, we remark that the eventual cost of the epidemic depends a great deal on the time horizon considered, and the value a society places on its elderly population (the group most at risk) (Keogh-Brown et al., 2009). Indeed, the metric used to evaluate a country’s economy is another factor that could change the calculation significantly (Marikina, 2018). These considerations are beyond the scope of the current paper, but provide interesting avenues of future work in this area, and would help countries prepare for future pandemics.

## 5 Conclusions

Our analysis suggests that the BC and federal governments were wise to impose severe shutdown levels at the beginning of the epidemic. The later reduction in shutdown levels was likely too large as compared to an optimal strategy. The difficulty is that people are unlikely to cooperate with rules that appear to be unnecessarily restrictive, especially when livelihoods are at stake (Bodas and Peleg, 2020; DiGiovanni et al., 2004). Future modelling studies that include behavioural components will be very helpful. Much work needs to be done by government and society to make sure that not only are countries economically prepared to handle future pandemics, but that populations also have the necessary understanding and mental resiliency to maintain a high level of prophylaxis for a long period of time.

## Data Availability

No data.

## Acknowledgements

This work was supported by (RCT) NSERC DG RGPIN-2016-05277 and (MTB) NSERC DG RGPIN-2016-03703. MTB initiated the project, and NDM wrote all of the code, ran the simulations, and wrote the first draft of the manuscript. RCT was part of the discussions throughout. MTB and RCT together wrote the final draft.

**Appendix A Slowly-varying SD is better when** *f* **is concave**

In Section 2.3 above, we stated that slowly-varying SD strategies would outperform rapidly-varying strategies because the shutdown effectiveness function *f* is concave. To see this relationship, consider a strategy with period 2*S*, where for the first half of each period *X*_*t*_ ≡ *x*_1_, and *X*_*t*_ ≡ *x*_2_ for the second half of each period. If the *x*_*i*_ are close enough to *x*_*c*_ so that the epidemic does not explode over a period of *S* days, then an analysis of the SEIQ equation shows that the effect of the shutdown is roughly linear, so that the periodic strategy above has approximately the same effect as a constant strategy *x*^*′*^ with 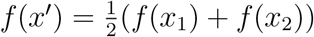. As *f* is concave,

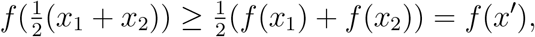

and so the periodic strategy is more expensive than a constant strategy with the same effectiveness. For example, if we take *X*_*t*_ ≡ *x*_*c*_ for the whole period of *T* days, the shutdown cost is $80.3b. If we alternate between values *x*_1_ = 0.28 and *x*_2_ = 0.09 (which satisfy 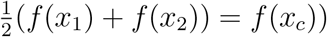 then the shutdown cost increases to $90.3b.

**Appendix B SD is non-zero**

In Section 2.2 we chose the shutdown effectiveness function *f* (*x*) = *x*^*θ*^, with *θ* < 1. We now show that this implies that the optimal strategy (with respect to total costs) is non-zero. For simplicity we just treat the case when we ignore excess deaths.

In a SEIR model the final population of susceptibles *S*_∞_ satisfies (Martcheva, 2015)

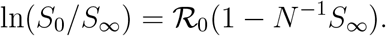

It follows that *S*_∞_ is a differentiable function of ℛ_0_: write *S*_∞_ = *H*(ℛ_0_). Let us write *S*_∞_ for the final number of susceptibles if we adopt a modified strategy *X*_*t*_ ≡ 0, and 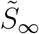 for the final number if *X*_*t*_ ≡ *x*, where *x* is small. Then

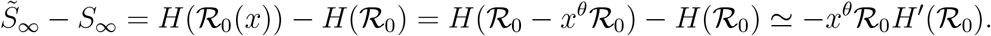

Hence the modified strategy leads to a cost saving (in terms of medical and deaths costs) of

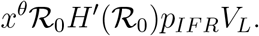

On the other hand, the economic cost of the modified strategy is *xg*_*D*_*T*, so writing *C*^(*T ot*^) and 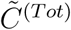 for the total costs of the original and modified strategies, we have

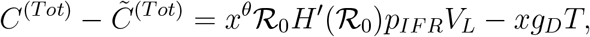

which is positive for small enough *x*.

A similar analysis shows that if we consider piecewise constant strategies, the optimal strategy *X*_*t*_ satisfies *X*_*t*_ > 0 in any time period in which the initial number of infected is greater than zero.

